# Somatic TP53 Mutations Drive T and NK Cell Dysfunction in AML and Can be Rescued by Reactivating Wild Type p53

**DOI:** 10.1101/2025.04.11.25325281

**Authors:** Li Li, Muharrem Muftuoglu, Edward Ayoub, Jiangxing Lv, Mahesh Basyal, Ran Zhao, Prashant S Menon, Aram Bidikian, Po Yee Mak, Yuki Nishida, Rupesh K Kesharwani, Ghayas C Issa, Navin Varadarajan, Bing Carter, Michael Andreeff

**Affiliations:** Section of Molecular Hematology and Therapy, Department of Leukemia, U.T. MD Anderson Cancer Center, Houston, Texas, USA; William A. Brookshire Department of Chemical and Biomolecular Engineering, University of Houston, Houston, Texas, USA; Department of Leukemia, U.T. MD Anderson Cancer Center, Houston, Texas, USA; Human Genome Sequencing Center, Baylor College of Medicine, Houston, Texas, USA

## Abstract

TP53-mutant acute myeloid leukemia (AML) is associated with particularly poor clinical outcomes and resistance to current therapies. While immunotherapy has revolutionized treatment for other hematologic malignancies, its efficacy in TP53-mutant AML remains limited. Although TP53 mutations in leukemic blasts are well characterized, their presence and functional consequences in immune cells have not been fully explored. Here we show that TP53 mutations are also present in T and NK cells from AML patients, where they are associated with increased activation markers but diminished cytotoxic function. T cells engineered to express common TP53 mutations exhibit an exhausted phenotype, characterized by impaired cytokine secretion and reduced tumor-killing capacity. Remarkably, restoring wild-type p53 conformation using a targeted small molecule reactivator reverses these dysfunctions and improves disease control in preclinical AML models. These findings reveal a novel mechanism of immune escape driven by mutant p53 in T cells and highlight the therapeutic potential of p53 reactivation. This work broadens our understanding of immune dysfunction in AML and supports incorporating immune-cell genotyping and correction strategies into future immunotherapy approaches for TP53-mutant disease.

## Main

The transcription factor p53, encoded by the TP53 gene, is pivotal as tumor suppressor, activated in response to cellular stressors such as DNA damage. This activation orchestrates the regulation of key genes involved in DNA repair, cell differentiation, cell cycle arrest, senescence and programmed cell death (apoptosis). Mutations in the TP53 gene are frequent in tumor cells, leading to diminished DNA binding capacity and, as a result, a reduced ability of p53 to act as tumor suppressor. In addition, p53 mutations can exert dominant-negative effect, further impairing any remaining functional p53^1^.

In Acute Myeloid Leukemia (AML) and myelodysplastic syndromes (MDS), patients with mutated TP53 (TP53m) carry on extremely poor prognosis ^2^, with intensive or targeted therapies, with a median overall survival rate of merely 6.5 months, in stark contrast to 33.6 months for individuals with wild-type TP53 AML^3^, depending established risk factors. These challenges underscore the critical need for novel and effective treatment strategies for TP53m AML, highlighting the imperative for innovative approaches in addressing this particularly resistant subset of AML.

While immunotherapy has significantly improved the outcomes for patients with lymphomas and myelomas and has even replaced chemotherapy in certain B-cell lymphoid malignancies^4^, the translation of these successes into the treatment of AML has been challenging^5^. The obstacles to successful immunotherapies in AML are linked to several factors, including genetic and clonal heterogeneity, a scarcity of viable leukemic targets, and an immunosuppressive tumor microenvironment^6,7^. In addition, little is known about mutations in immune cells. Previous studies^8,9^ have identified mutations in T-cells, such as TET2, IDH1/2, and DNMT3a, primarily considering these as indicators of clonal hematopoiesis rather than an key contributor to leukemia pathobiology or potential therapeutic targets.

The efficacy of immunotherapies and to a degree of chemo-and targeted therapies largely depends on the functional response of immune cells, including the ability of T cells and NK cells to eliminate cancer cells. Investigating the mutations in these immune cells could unveil novel insights into the mechanisms by which they impair the immune system’s capacity to combat cancer, potentially unveiling novel therapeutic strategies to enhance the efficacy of immunotherapy in AML patients.

In this study, we discovered p53 mutations in T cells and investigated the impact of those on anti-tumor function and fitness of immune cells, in AML. To our knowledge, this is the first report exploring the functional consequences of p53 mutations on immune cells from AML patients and to develop a novel therapeutic strategy to restore T cell functionality. Our findings reveal a new mechanism by which p53 mutations impair immune cell function against AML cells, potentially explaining the failure of immunotherapies in these patients.

## Results

### T and NK cells exhaustion is associated with TP53 mutations in AML patients

AML is a heterogeneous bone marrow disorder resulting from the accumulation of complex mutations in hematopoietic stem cells (HSCs). While many therapies target various mutations in AML, TP53 mutations remain without any effective targeted treatments, and their impact on immune cells is still largely unknown. To address this, we employed single-cell technologies, including transcriptomics, proteomics, and secretomics, to dissect the bone marrow and its microenvironment from AML patients and comprehensively analyze the impact of p53 mutations on immune cells.

In our previous study ^10^, we demonstrated that T-cells from AML patients harboring TP53 mutations exhibit impaired polyfunctionality, as evidenced by altered cytokine secretion profiles. To obtain a deeper understanding of the immune-cell landscape in TP53-mutant AML, we performed single-cell RNA sequencing and employed an unsupervised analytical approach to investigate transcriptional changes at the single-cell level of immune populations, specifically T and NK cells. For an unbiased comparison across patients, we pooled scRNA-seq data from the bone marrow of 24 newly diagnosed AML patients for downstream analysis. Dimensional reduction using Uniform Manifold Approximation and Projection (UMAP) (**Fig. 1A**) enabled the visualization of distinct immune cell clusters, while unsupervised clustering identified cell types based on lineage-specific gene expression^11^. To assess T and NK cell functionality, we calculated an exhaustion score, defined as the mean expression of 21 exhaustion-associated markers (**Supplementary Table 1**). Notably, this analysis revealed significantly elevated exhaustion scores in T and NK cells from TP53-mutant AML patients compared to their TP53 wild-type counterparts (**Fig.1B**). Increased expression of canonical exhaustion markers, including TIGIT, PD-1, LAG-3, and TIM-3, was particularly evident in TP53-mutant samples (**Fig. 1C**), suggesting a pronounced exhaustion phenotype. We further assessed the protein-level expression of exhaustion markers PD-1, TIGIT, and TIM-3 using CyTOF analysis, which enables high-parametric single-cell protein quantification. To minimize the batch effect, we employed live-cell barcoding approach^12^, allowing sample pooling after surface staining while preserving individual sample identity. A total of 38 AML bone marrow (BM) samples were analyzed, comprising 26 TP53-mutant (TP53m) and 12 TP53 wild-type (TP53wt) AML cases, along with 13 TP53wt chronic myelomonocytic leukemia (CMML) BM samples and four BM samples from healthy donors. CyTOF analysis confirmed the significant upregulation of PD-1 and TIGIT in CD3+ T cells from TP53m compared to TP53wt AML patients. Furthermore, NK cells from TP53m AML patients also exhibited increased expression of PD-1, TIGIT, and TIM-3 relative to both TP53wt AML patients and healthy donors (**Fig. 1D**), further corroborating the transcriptional evidence of immune exhaustion. To assess the functional implications of these findings, we evaluated cytokine secretion capacity using a single-cell fluorescence-based ELISA assay (Isoplexis). This assay quantified the secretion of 32 cytokines at the single-cell level following stimulation with anti-CD3/CD28 microbeads. Analysis revealed a significant reduction in the polyfunctional strength index (PSI) in both CD4+ and CD8+ T cells from TP53m AML patients, indicating impaired effector function. Notably, the decrease in PSI was driven primarily by reductions in granzyme B, IFN-γ, MIP-1α, perforin, and TNF-α, key mediators of cytotoxic T-cell responses (**Fig. 1E**). Collectively, these findings demonstrate that T and NK cells from TP53-mutant AML patients exhibit greatly increased exhaustion at both the transcriptional and functional levels, providing further insights into the immune dysfunction associated with TP53 mutations in AML.

**Figure 1.**
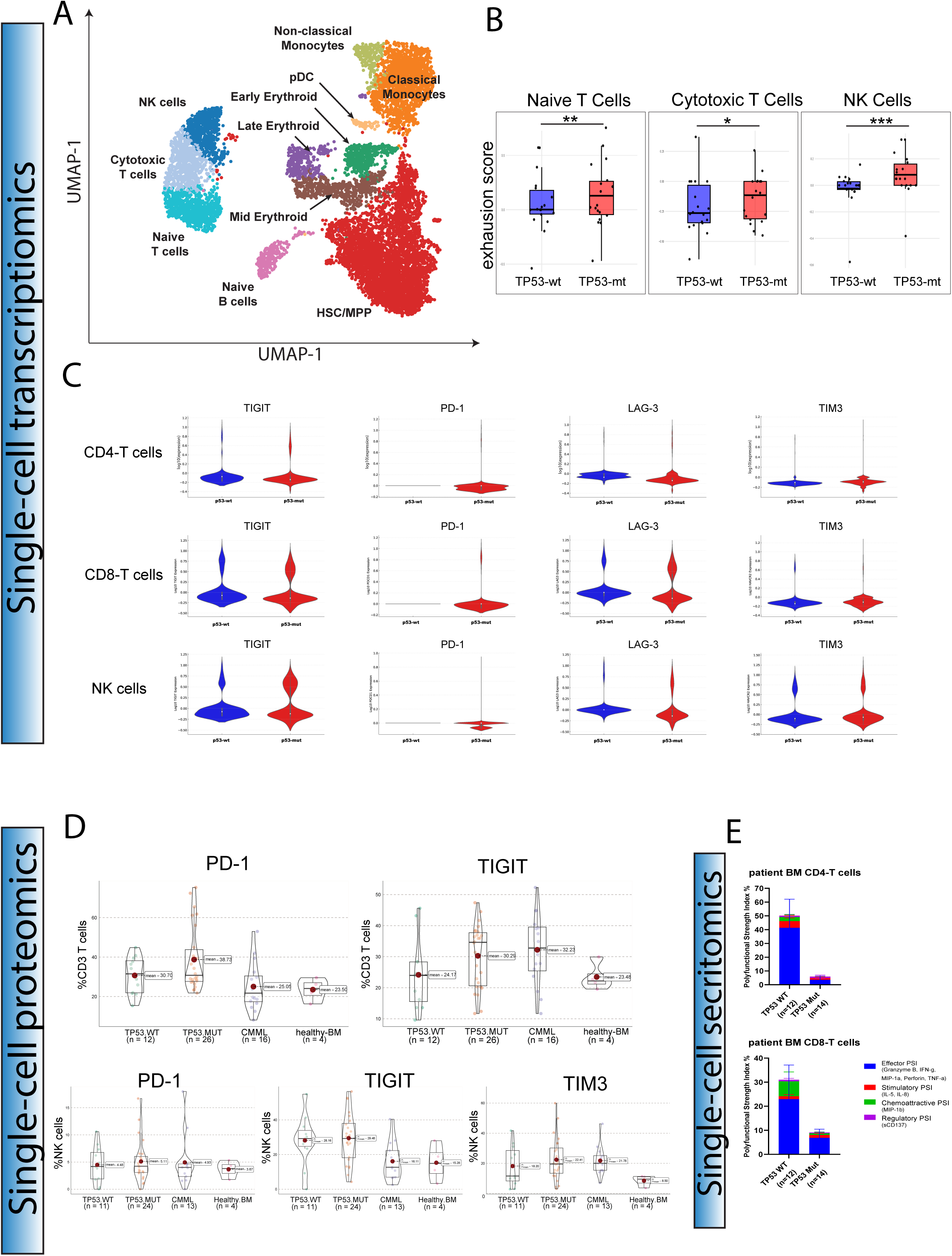
T and NK cells exhaustion is associated with TP53 mutations in AML patients.

### TP53 mutations occur in T and NK cells in AML patients and alter their phenotype

While TP53 mutations in AML patients have been associated with altered immune response^6,7^, only few studies^8,9^ have identified specifically addressed the presence of TP53 mutations in immune cells. Using digital droplet PCR(ddPCR), we demonstrated that TP53 mutations in AML patients arise somatically in T cells, independent of Li-Fraumeni Syndrome (LFS). In this study, we utilized whole-exome sequencing (WES) to detect TP53 mutations in AML patients and confirmed these mutations in highly purified (>99%) CD3+CD34− FACS-sorted cells using digital droplet PCR (ddPCR).Our results revealed that TP53 mutations were present in over 20% of droplets, and the mutations identified in T cells were identical to those detected in leukemic blasts^13^ (Fig. 2A). Simultaneously, we performed single-cell DNA sequencing integrated with surface antigen expression analysis, enabling direct phenotype-genotype correlations from bone marrow (BM) samples of four AML patients with TP53 mutations detected by WES. Cells were clustered based on the expression of 45 oligo-conjugated antibodies and are colored by cell types. Using UMAP analysis, we visualized various cell types identified by surface antigen expression, identifying T cells as CD3-positive and NK cells as CD56-positive. Notably, by integrating single-cell DNA sequencing data with UMAP clustering, we found that TP53 mutations were widely distributed across different immune cell lineages. These mutations were observed not only in myeloid lineages (e.g., monocytes and leukemic cells) but also in lymphoid cell subsets, including T, B, and NK cells. Importantly, TP53 mutations were detected in subsets of T and NK cells across all four AML patient samples, with more than 95% of mutations being monoallelic. (**Fig. 2B**). At the single-cell resolution level, we quantified the frequency of TP53 mutant T and NK cells across four AML patients. TP53 mutations were detected in CD4, CD8 T, and NK cells, with mean mutation frequencies of 32% in CD8 T cells (range: 7%–91%), 36% in CD4 T cells (range: 4%–89%), and 32% in NK cells (range: 9%–86%) (**Fig. 2C**). To further characterize the phenotypic impact of TP53 mutations on T and NK cells, we conducted a comprehensive analysis of surface antigen expression using a heatmap. Distinct phenotypic profiles were observed across different immune cell subsets, with CD4 and CD8 T cells displaying high CD5 and CD7 expression, while NK cells exhibited high CD16 expression, as expected. Notably, TP53mut T and NK cells displayed phenotypic alterations compared to their TP53 wild-type counterparts (**Fig. 2D**). To further explore these differences, we performed a comparative analysis of TP53mut and TP53wt T and NK cells. We applied unsupervised clustering to analyze phenotypic differences, incorporating TP53 mutation status. Interestingly, CD4, CD8, and NK cells were distinctly separated in the UMAP, and TP53mt T and NK cells formed unique clusters, further underscoring their distinct phenotypic profiles (**Fig. 2E**). Indeed, we observed differential expression of multiple immune-related markers between TP53mt and TP53wt T and NK cells from the same patient sample. Specifically, CD69, CD71, and CD38 were significantly increased in TP53mt CD4 and CD8 T cells compared to their TP53wt counterparts, suggesting enhanced proliferative capacity in the presence of TP53 mutations. More importantly, CD2 expression, a key molecule for immunological synapse formation and T-cell anti-tumor function^14^, was markedly reduced in TP53^mt^ CD4 and CD8 T cells, suggesting a potential impairment in T-cell functionality. Similarly, TP53 mutant NK cells exhibited elevated expression of activation markers such as CD44, CD69, and CD71 (**Fig. 2F**), indicating a TP53 mutation-driven activation phenotype. These findings confirm that TP53 mutations occur in immune cells, including T and NK cells, and profoundly reshape the immune landscape in AML patients. While TP53 mutations enhance the proliferative and activation profiles of T and NK cells, they simultaneously impair T-cell functionality, potentially contributing to T-cell exhaustion. Based on these data, we hypothesize that monoallelic TP53 mutations impair normal p53 function in T cells, which is essential for cell cycle regulation and proliferation control. Through a dominant-negative effect, these mutations may drive T-cell exhaustion and dysfunction, ultimately compromising anti-leukemic immune responses.

**Figure 2.**
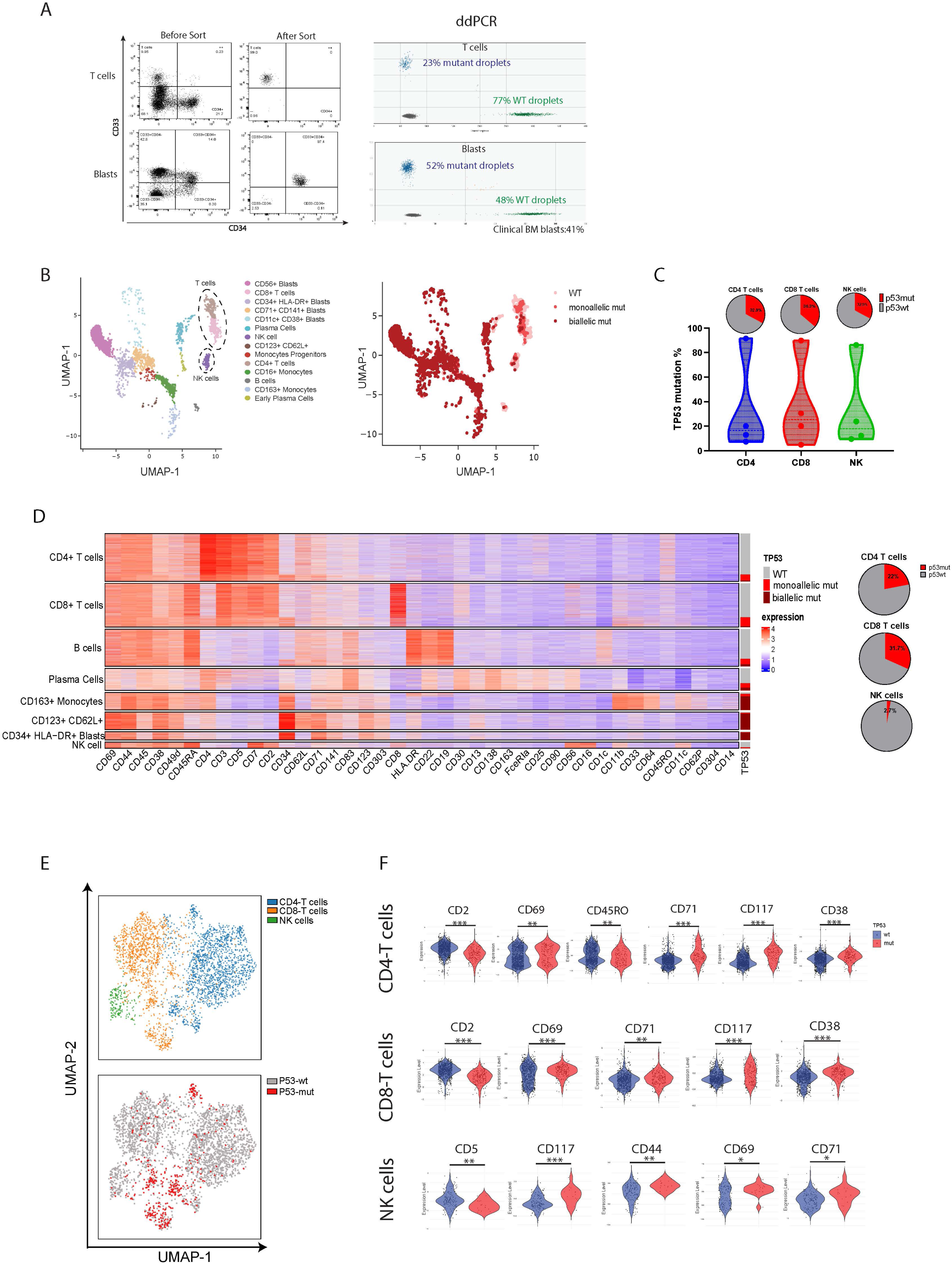
TP53 mutations occur in T and NK cells in AML patients and alter their phenotype.

### Impact of Mutant p53 in CAR-T cells phenotype

To test our hypothesis, we engineered CD123-targeted CAR-T cells overexpressing the mutant p53-Y220C using a lentiviral vector to specifically target AML cells. This specific p53 mutation was chosen because of its reversibility by PC14586, a small molecule that could restore mutant p53 to its wild-type conformation and reactivate the functions of WT p53 protein. The overexpressed mutant p53 is expected to exert a dominant-negative effect on wild-type p53. Control CAR-T cells were generated without overexpression of mutant p53, serving as a baseline comparison (**Fig. 3A**). Notably, when we overexpressed wild-type p53, these T cells underwent apoptosis. Prior to assessing the functionality of the p53mt CAR-T cells, we confirmed the successful transduction of the engineered CAR construct into T cells. Utilizing a specific CAR antibody, we observed CAR expression on over 85% of the transduced T cells. Remarkably, p53 protein was detected in 80% of the p53mt CAR-T cells, while p53 protein was detected at low level (<5%) in the control CAR-T cells, as expected (**Fig.3B**). Subsequently, we tested the autonomous growth of p53mt CAR-T cells in comparison to control CAR-T cells. We observed that p53mt CAR-T cells proliferated more rapidly than control CAR-T cells under normoxic (5% CO2 at 37°C) as well as hypoxic conditions (1% oxygen at 37°C) for 7 days (**Fig. 3C**).

**Figure 3.**
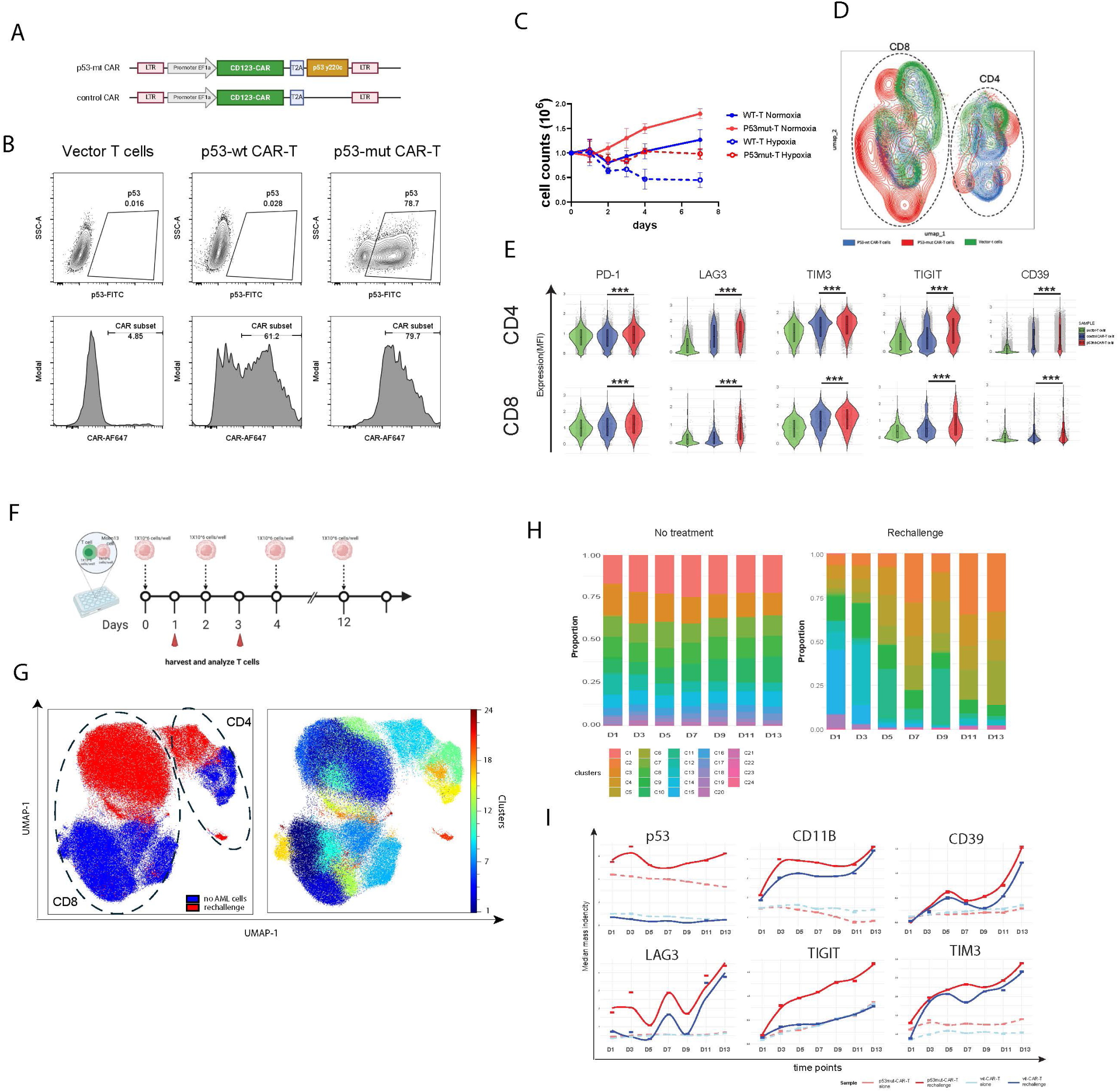
Impact of Mutant p53 in CAR-T cell phenotype

These results suggest that mutant p53 enhances the proliferative capacity of CAR-T cells and enable them to overcome hypoxic conditions. Next, we characterized the phenotypic profile of the CAR-T cells using CyTOF analysis using a 50-parameter T-cell-focused panel. UMAP analysis revealed that p53 mutant (p53mt) CAR-T cells exhibited a distinct profile compared to both control CAR-T cells and empty vector T cells (V-T cells), particularly within the CD4 T-cells subset (**Fig. 3D**). To further investigate our hypothesis that mutant p53 induces T-cell exhaustion, we assessed the expression of exhaustion markers on p53mt CAR-T cells relative to control CAR-T cells. Notably, the expression levels of PD-1, LAG-3, TIM-3, TIGIT, and CD39 were significantly increased in both CD4 and CD8 p53mt CAR-T cells compared to control CAR-T cells(**Fig.3E**). These results demonstrated that mutant p53 not only enhances the proliferative capacity of CAR-T cells but also induces a pronounced state of T-cell exhaustion.

To investigate the impact of CAR-T cells upon repeated antigenic exposure, we developed an in vitro rechallenge model. In this system, p53mt CAR-T cells were co-cultured with CD123-positive Molm13 cells at a 1:1 ratio in the absence of exogenous cytokine support. To mimic sustained antigen exposure, fresh Molm13 cells were replenished every 48 hours. which lacked mutant p53 expression and served as a baseline comparator, underwent identical conditions (**Fig.3F**).

To characterize the dynamic functional and phenotypic changes in p53mt CAR-T cells during AML interactions, cells were collected from the co-culture every two days for 14 days and analyzed by CyTOF using a previously described live-cell barcoding approach^12^(**Supplementary Table 2**). By day 14, cytotoxicity assays revealed that control CAR-T cells efficiently eradicated over 99% of Molm13 cells, while p53mt CAR-T cells exhibited markedly reduced cytotoxic activity, with 11% of Molm13 cells persisting in the culture. This significant impairment in cytolytic function under repeated antigenic stimulation suggests that mutant p53 compromises CAR-T cell effector capabilities, potentially through exhaustion-mediated dysfunction. Further analysis using UMAP clustering revealed that rechallenged p53mt CAR-T cells displayed a distinct proteomic signature compared to their unstimulated counterparts, indicative of chronic antigenic engagement triggering extensive proteomic reprogramming via CAR signaling (**Fig. 3G**). This shift suggests that prolonged AML exposure alters key functional states within p53mt CAR-T cells, likely impacting their persistence and therapeutic efficacy.

To further delineate these proteomic changes, we performed Phenograph clustering analysis, a computational approach for resolving subpopulations within high-dimensional single-cell data. Across 28 CAR-T cell samples collected at seven different time points, we identified 24 distinct clusters (**Fig. 3H**). The proportions of these clusters remained stable in unstimulated p53mt CAR-T cells but exhibited notable shifts following antigenic rechallenge. Specifically, cluster 2 expanded from 2% on day 1 to 32% on day 13, characterized by elevated expression of exhaustion markers PD-1, TIGIT, TIM-3, and LAG-3. Conversely, cluster 15, enriched for high expression of Eomes and T-bet, key regulators of T-cell memory and effector function^15^, declined from 28% on day 1 to 0.2% on day 13, suggesting a substantial loss of memory-like and cytotoxic potential in p53mt CAR-T cells. This decline in memory-associated transcription factors likely contributes to impaired persistence and reduced long-term efficacy, exacerbating exhaustion-driven dysfunction. Additionally, CD27, a crucial marker for T-cell survival and activation^16^, was markedly downregulated (**Fig.3H, supplementary Fig. 1**). To assess the temporal evolution of exhaustion marker expression, we examined p53 levels over time, as its dysregulation has been implicated in T-cell exhaustion and impaired persistence. Tracking p53 expression dynamics allows us to determine whether its accumulation contributes directly to the upregulation of inhibitory receptors, thereby influencing CAR-T cell efficacy. In rechallenged p53mt CAR-T cells, p53 expression progressively increased, whereas no such trend was observed in unstimulated p53mt CAR-T cells. As expected, p53 expression remained undetectable in control CAR-T cells, irrespective of antigen exposure. Exhaustion markers, including CD39, LAG-3, TIGIT, and TIM-3, were upregulated in both p53mt and control CAR-T cells upon repeated antigen stimulation. However, p53mt CAR-T cells exhibited an accelerated and heightened expression of these exhaustion markers compared to controls, potentially driven by aberrant activation of NF-κB and STAT3 signaling pathways. Notably, TIGIT expression demonstrated a pronounced increase, reaching twice the level observed in control CAR-T cells by the final time point. The mechanistic link between mutant p53 expression and TIGIT upregulation remains under investigation (Fig. 3I). Collectively, these findings suggest that mutant p53 accelerates the acquisition of exhaustion markers, rendering p53mt CAR-T cells more prone to functional exhaustion upon sustained antigenic stimulation, thereby compromising their anti-tumor efficacy.

### Mutant p53 impairs CAR-T cell anti-tumor function in vitro

Based on our phenotypic observations, we hypothesized that mutant p53 impairs the anti-tumor function of CAR-T cells. To test this hypothesis, we employed the Incucyte Live-Cell Analysis System to evaluate the cytotoxic capacity of p53mt CAR-T cells against their target cells by continuously tracking the number of apoptotic and surviving target cells in real time. This real-time monitoring approach provides a more dynamic and precise assessment of cytotoxicity compared to traditional endpoint assays, allowing for a detailed understanding of temporal killing kinetics and functional differences between p53mt and control CAR-T cells. p53mt CAR-T cells exhibited a significant reduction in cytotoxicity compared to control CAR-T cells (Fig. 4A), suggesting that the anti-tumor function of p53mt CAR-T cells is compromised due to mutant p53 expression. Moreover, we conducted a single-cell cytokine secretion assay to assess the polyfunctionality of these cells under stimulation. The data revealed a marked reduction in the polyfunctional strength index (PSI) in both CD4 and CD8 p53mt CAR-T cells compared to control CAR-T cells, particularly for effector cytokines such as Granzyme B, IFN-γ, MIP-1α, perforin, and TNF-α, as well as IL-5 and IL-8, which are associated with T-cell activation (Fig. 4B). Notably, this cytokine downregulation aligns with well-established exhaustion signatures observed in dysfunctional CAR-T cells^17^, suggesting that mutant p53 may drive an early onset of exhaustion through altered transcriptional or metabolic pathways. Additionally, we determined the production of TNF-α, IFN-γ, and IL-2 in p53mt CAR-T cells using a flow cytometry-based approach. The results confirmed that p53mt CAR-T cells produced lower levels of these cytokines compared to control CAR-T cells (Fig. 4C). These findings collectively indicate that the presence of mutant p53 not only impairs the cytotoxic function of CAR-T cells but also diminishes their cytokine secretion capacity, ultimately leading to reduced anti-tumor efficacy. To determine whether this phenomenon is specific to the p53-Y220C mutation or a broader effect of p53 mutations, we also engineered CAR-T cells with the p53-R175H mutation. These cells exhibited the same exhaustion-associated features and reduced cytotoxicity (**supplementary Fig. 2**). Mechanistically, this could be attributed to transcriptional repression via NF-κB, which is known to regulate cytokine expression^18^, or metabolic constraints imposed by mutant p53, limiting energy availability for cytokine production. Further studies are required to dissect these pathways and their contribution to CAR-T cell dysfunction.

**Figure 4.**
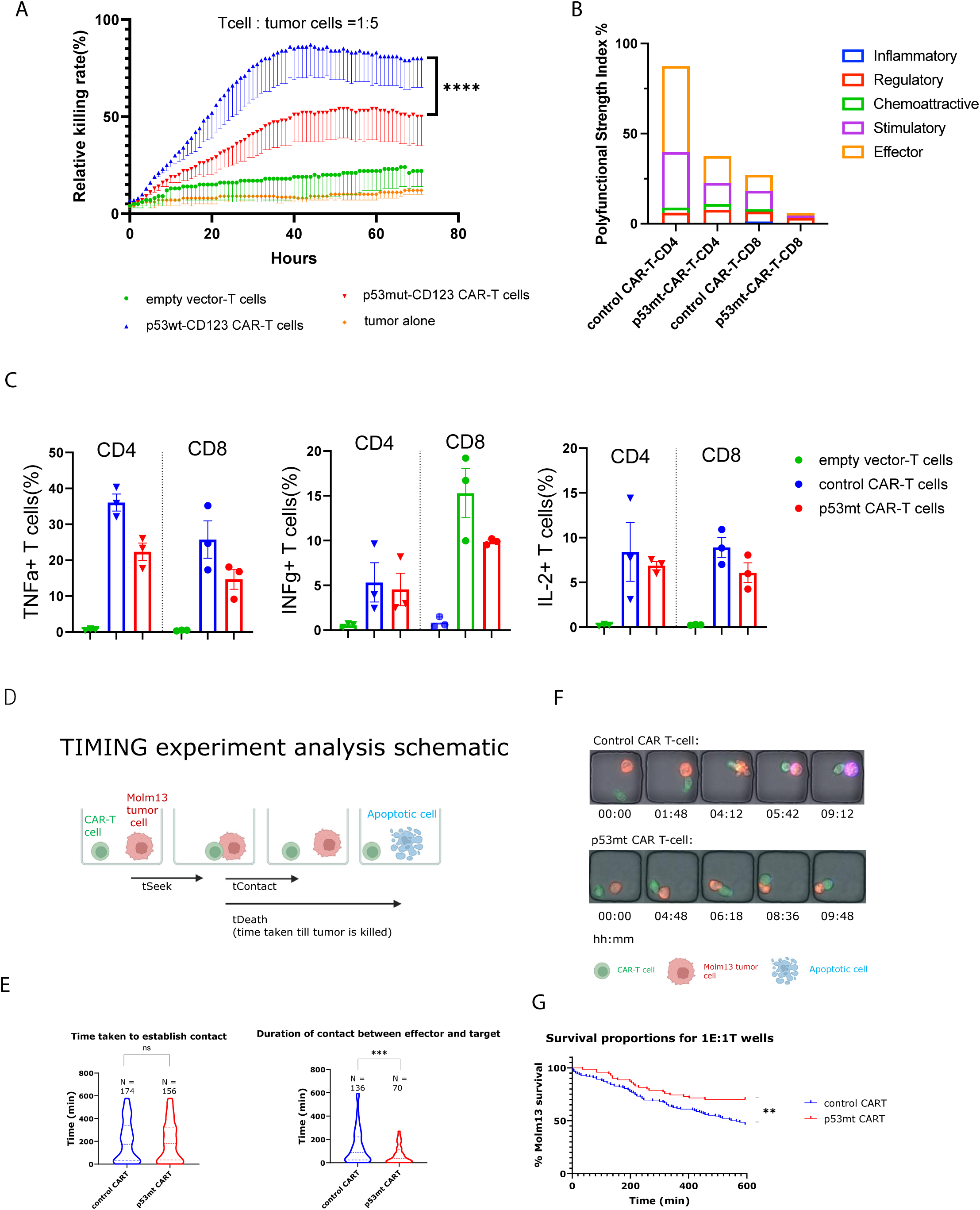
Mutant p53 impairs CAR-T cells anti-tumor functoin in vitro

Additionally, we employed Timelapse Imaging Microscopy in Nanowell Grids (TIMING) to assess the functionality of p53 mutant (p53mt) CAR-T cells compared to control CAR-T cells, thereby dissecting functional heterogeneity at the single-cell level. TIMING offers a distinct advantage over traditional imaging or cytotoxicity assessment methods by enabling high-resolution, real-time tracking of individual cell interactions. This approach provides detailed insights into CAR-T cell engagement, synapse formation, and killing efficiency, allowing for a more precise evaluation of functional impairments in p53mt CAR-T cells. The cytotoxic process was delineated into three distinct phases: the time taken for CAR-T cells to seek out tumor cells (tSeek), the duration of contact between CAR-T cells and tumor cells (tContact), and the time required to induce tumor cell apoptosis (tDeath), at an effector-to-target (E) ratio of 1:1 (Fig. 4D). Interestingly, the time required for CAR-T cells to establish contact with tumor cells (tSeek) did not differ significantly between p53mt and control CAR-T cells (Fig. 4E). However, the duration of contact between effector and target cells (tContact) was significantly shorter for p53mt CAR-T cells compared to control CAR-T cells, suggesting that p53mt CAR-T cells exhibit impaired synapse formation ability (Fig. 4F). This impairment in synapse formation is consistent with the observed decreased propensity of p53mt CAR-T cells to eliminate AML cells. This defect may be attributed to the downregulation of adhesion molecules, such as CD2, which were observed in AML patients samples(Fig.2). Furthermore, the survival proportions of Molm13 tumor cells in 1:1 effector-to-target (E) ratio over time, as shown in the survival curves, indicated that Molm13 cells had a higher survival rate when co-cultured with p53mt CAR-T cells compared to control CAR-T cells. This increased survival could result from reduced cytolytic function, impaired tumor recognition, or a combination of both factors. Defects in immune synapse formation, diminished cytokine secretion, or altered metabolic fitness may contribute to the overall functional impairment of p53mt CAR-T cells. This finding further underscores the impaired cytotoxic function of p53mt CAR-T cells (Fig. 4G, supplementary video). Taken together, all in vitro data demonstrate that mutant p53 impairs the anti-tumor function of CAR-T cells by reducing cytokine secretion and downregulating synapse formation.

### Mutant p53 impairs CAR-T cell eliminate AML cells in vivo

To investigate the in vivo impact of mutant p53 on CAR-T cell functionality, we generated a patient-derived xenograft (PDX) mouse model using venetoclax-resistant AML patient cells, as described in the Methods section. Briefly, mice were intravenously injected with luciferase-transduced PDX cells. After confirming the engraftment of PDX cells by detecting 1% human CD45 in mouse blood, we initiated treatment with p53 mutant (p53mt) CAR-T cells, control CAR-T cells, or empty vector T cells via tail vein injection. Tumor burden was monitored using bioluminescence imaging (BLI) and overall survival was assessed. The BLI data revealed that mice treated with p53mt CAR-T cells exhibited an increased AML burden by day 47 post-treatment. In contrast, mice treated with control CAR-T cells maintained the suppression of AML cells until day 120 post-treatment(Fig.5B). The BLI signals indicated a significantly higher tumor burden in mice treated with p53mt CAR-T cells compared to those treated with control CAR-T cells and empty vector T cells, as well as untreated PDX mice. This increased tumor burden may correlate with reduced CAR-T cell persistence or expansion in vivo, potentially due to the exhaustion phenotype observed in vitro. Further analysis of CAR-T cell engraftment, proliferation, and exhaustion marker expression in treated mice is necessary to gain deeper insights into the mechanisms underlying this impaired anti-tumor response.

**Figure 5.**
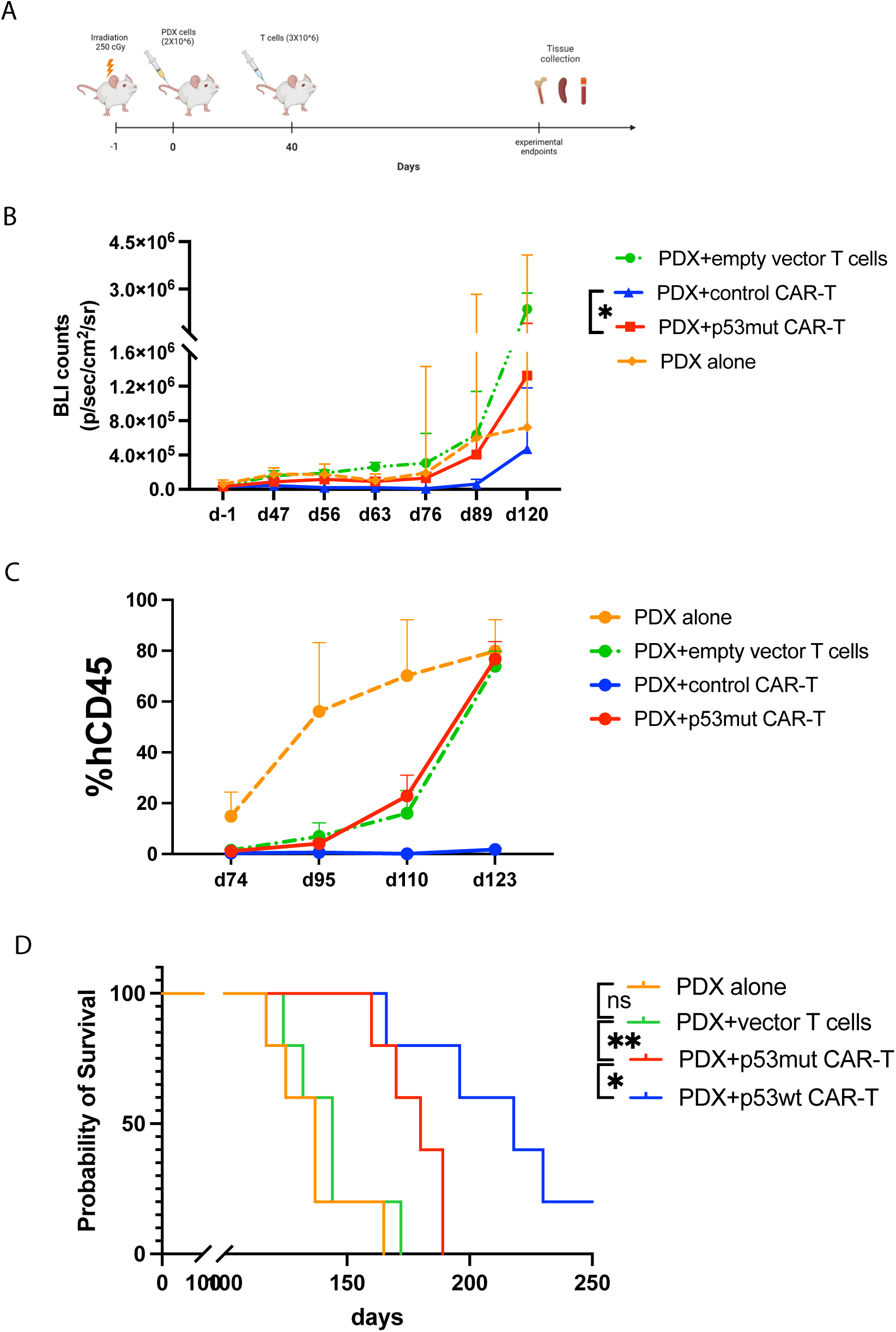
Mutant p53 impairs CAR-T cells eliminate AML cellsin vivo

To validate the BLI imaging findings and evaluate systemic tumor growth, we quantified circulating AML cells by measuring human CD45 (hCD45) levels using flow cytometry in mouse blood at various time points. Consistent with the BLI data, no human cells were detectable in the blood of mice treated with control CAR-T cells. However, mice treated with p53mt CAR-T cells showed a marked increase in circulating AML cells, with over 70% hCD45+ cells detected by day 123 post-treatment (Fig.5C). This was in stark contrast to the control CAR-T cell-treated group, which had undetectable levels of hCD45+ cells. Crucially, survival analysis revealed that mice treated with p53mt CAR-T cells exhibited significantly worse survival outcomes compared to those treated with control CAR-T cells (Fig.5D). Consistent with the exhaustion phenotype observed in vitro, CARl7JT cells harvested from the bone marrow of p53mutl7JCAR-T cells treated mice expressed markedly higher levels of PDl7J1, TIMl7J3, TIGIT, LAGl7J3, CD39, and CTLAl7J4 than CARl7JT cells from control animals (**Supplementary Fig. 3**).These findings collectively suggest that p53mt CAR-T cells are significantly less effective in controlling tumor growth compared to control CAR-T cells, highlighting the detrimental effects of mutant p53 on CAR-T cell efficacy in vivo.

### Reactivating Mutant p53 in T Cells Enhances Anti-Tumor Activity

Based on our phenotypic and functional observations that mutant p53 drives T-cell exhaustion and impairs their anti-tumor function, we hypothesized that correcting the structure of mutant p53 might restore T-cell functionality. Mechanistically, mutant p53 may contribute to exhaustion by dysregulating key transcriptional programs involved in T-cell proliferation. To test this hypothesis, we utilized a small molecule, PC14586, which has been reported to fill the structural gap in mutant p53-Y220C protein, thereby restoring its conformation and transcriptional activity to that of the wild-type and reactivating its function. This molecule is currently under investigation in clinical trials^19^, as well as in preclinical and most recently in a clinical trial in AML ^20^.

To evaluate the efficacy of the p53 reactivator in T cells, we first treated our p53mt CAR-T cells in vitro with the recommended dose (8 μM) added to the culture media. We then assessed the expression levels of the mutant p53 protein in samples with and without the reactivator using a specific antibody that recognizes the mutant p53 protein. As expected, both CD4 and CD8 T-cells exhibited reduced levels of mutant p53 in p53mt CAR-T cells following treatment with the p53 reactivator (Fig. 6A). The reduction in mutant p53 protein levels time-dependent, with a 50% reduction in CD4 T-cells and a 38% reduction in CD8 T-cells observed at 72 hours (Fig. 6B). This finding was confirmed by Western blot analysis (**Supplementary Fig. 4**), which showed increased expression of p21 and MDM2 following p53 reactivator treatment, suggesting that the reactivator either facilitates the degradation of misfolded mutant p53 or promotes its refolding into a wild-type-like conformation.

**Figure 6.**
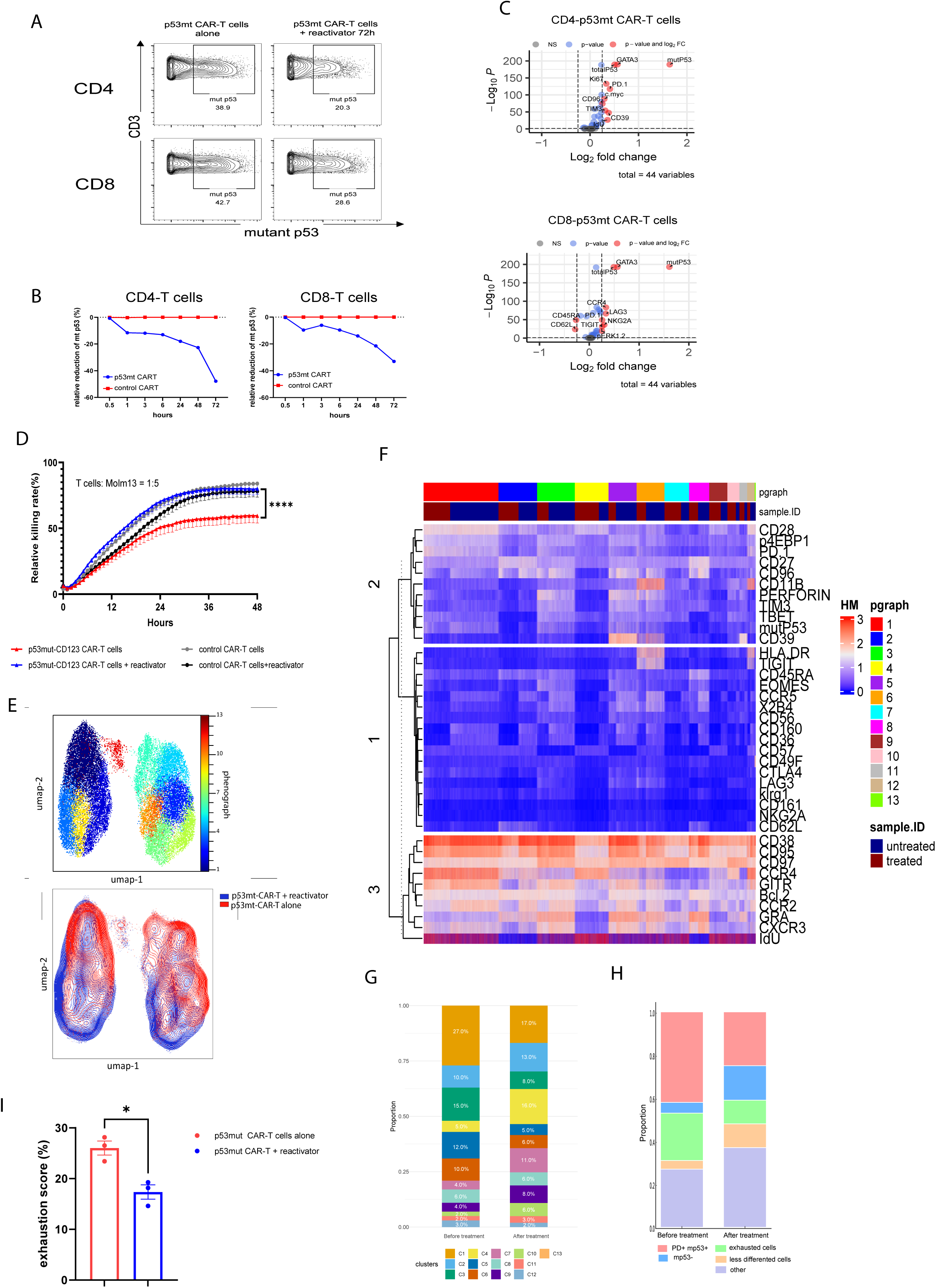
Reactivating Mutant p53 in T Cells Enhances Anti-Tumor Activity.

To further investigate the impact of mutant p53 on T-cell phenotype, we conducted a differential expression analysis comparing mutant p53-expressing T-cell subsets to those not expressing the mutant protein. Interestingly, T-cells expressing mutant p53 exhibited significantly increased levels of exhaustion markers: PD-1, TIM-3, and CD39 in CD4 T-cells, and LAG-3, TIGIT, and PD-1 in CD8 T-cells (**Fig. 6C**). Additionally, there was a significant reduction in the expression of naïve memory markers, CD45RA and CD62L, in CD8 T-cells. These findings corroborate our previous observations that mutant p53 induces T-cell exhaustion and terminal differentiation. To further investigate the functional impact of p53 reactivation on CAR-T cells, we performed a real-time cell killing assay using the InCuCyte system. The data demonstrated that p53 reactivator-treated p53mt CAR-T cells exhibited improved cytotoxic capacity **(Fig. 6D**), confirming that p53 reactivation enhances the anti-AML efficacy of CAR-T cells.

To explore the mechanisms underlying the functional restoration of p53mt CAR-T cells following p53 reactivator treatment, we conducted CyTOF analysis. Using high-dimensional reduction analysis via UMAP, we visualized cellular similarities on a two-dimensional map. The results demonstrated that p53mt CAR-T cells treated with the reactivator clustered predominantly in the top right of the map, while untreated cells were located in the bottom left, with some overlap between the two groups (**Fig. 6E**). This spatial distribution suggests a significant alteration in the proteomic landscape of p53mt CAR-T cells post-reactivator treatment. To further characterize the subpopulations within the CD4 and CD8 T-cell compartments, we employed the unsupervised clustering algorithm Phenograph, which identified 13 distinct clusters within the entire T-cell population (**Fig.6E**). Heatmap analysis revealed that each cluster exhibited a unique profile, and the proportions of these clusters were markedly altered following p53 reactivator treatment (**Fig. 6 F,G**). Notably, the proportions of clusters 1, 3, and 5 were significantly reduced upon treatment. Phenotypic analysis of these clusters indicated that they were enriched for mutant p53-positive subsets.

To gain deeper insights into the landscape changes, we consolidated certain clusters into five meta-clusters based on their shared phenotypic characteristics. This strategy facilitated a more comprehensive visualization of the modifications driven by p53 reactivator treatment. Meta-cluster 1, defined by elevated PD-1 and mutant p53 expression, declined from 43% to 25%. Meta-cluster 3, enriched in exhaustion markers including TIGIT, TIM-3, LAG-3, and CD39, decreased from 22% to 11%. Conversely, meta-cluster 2, lacking mutant p53 expression, expanded from 5% to 16%. Furthermore, meta-cluster 7, representing a less differentiated T-cell subset, grew from 4% to 11% (**Fig. 6H**). These findings were substantiated by the exhaustion score, as detailed in the Methods section, which revealed a significant reduction in exhaustion levels in p53mt CAR-T cells following p53 reactivator treatment (**Fig. 6I**). Taken together, these data indicate that the p53 reactivator rescues the anti-tumor efficacy of p53mt CAR-T cells by correcting mutant p53, alleviating exhaustion, and optimizing their phenotypic and functional properties, ultimately enhancing their therapeutic potential against AML.

### Reactivation of Mutant p53 in CAR-T Cells Prolongs Mouse Survival

To further evaluate the in vivo efficacy of the p53 reactivator in restoring the anti-tumor function of p53 mutant (p53mt) CAR-T cells, we employed our venetoclax-resistant patient-derived xenograft (PDX) mouse model. Mice received a single intravenous dose of 3 × 10^6 T cells expressing anti-CD123 CAR and mutant p53 pre-treated with the p53 reactivator. Control groups included T cells expressing anti-CD123 CAR with or without mutant p53, or empty vector-transduced T cells, administered on day 1 following confirmation of engraftment in mouse blood (**Fig. 7A**). Mouse survival and circulating AML cells were monitored via flow cytometry. Human cells in the blood were identified using anti-human CD45 antibodies, followed by CD3 and CD33 staining to distinguish T cells from AML cells (**Fig. 7B**).

**Figure 7.**
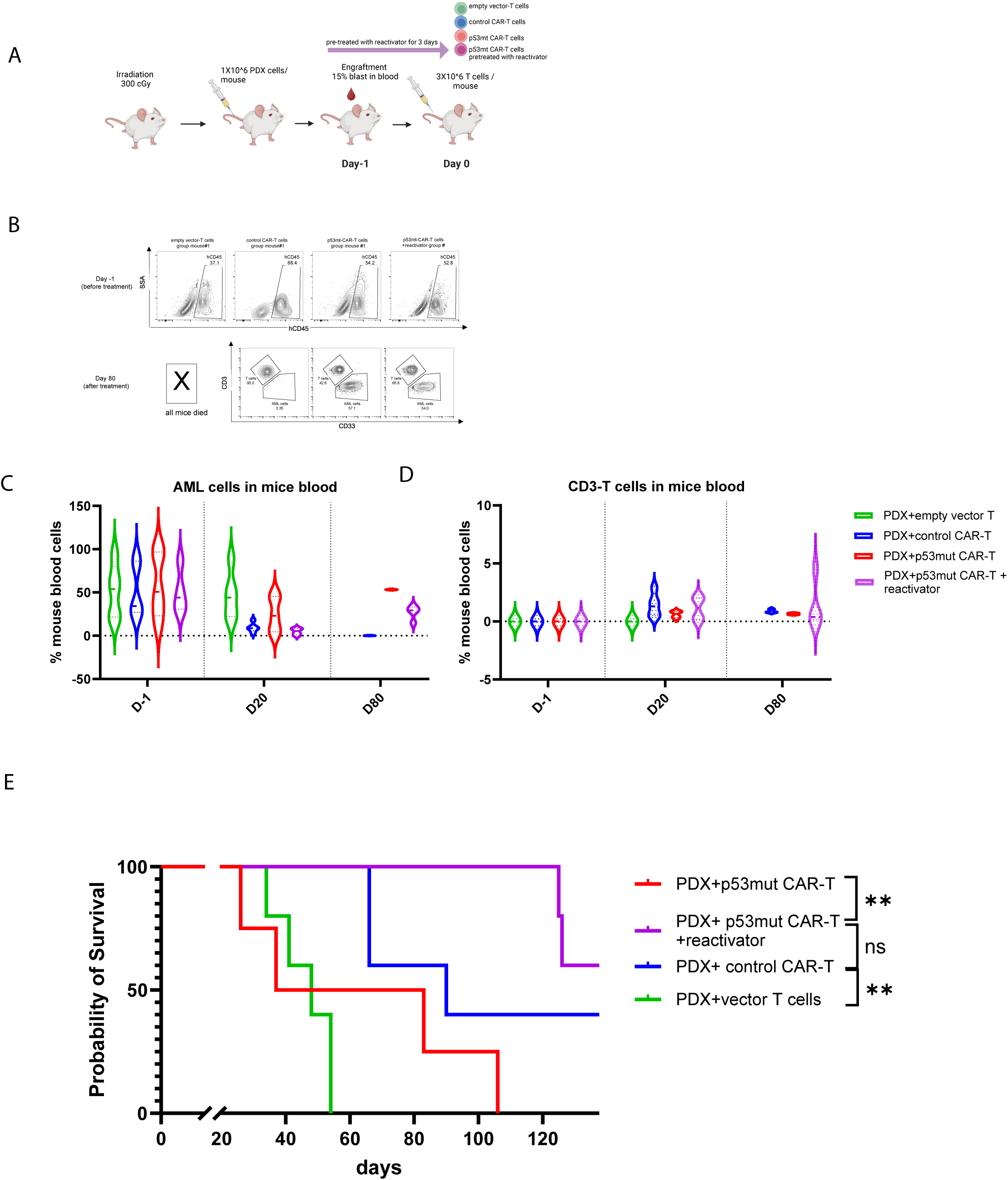
Reactivation of Mutant p53 in CAR-T cells prolongs mouse survival

Prior to T-cell administration, AML cell distribution was consistent across all groups. By day 20 post-treatment, AML cells were effectively eradicated in mice receiving reactivator pre-treated p53mt CAR-T cells, as well as those treated with control CAR-T cells. In contrast, mice receiving untreated p53mt CAR-T cells exhibited persistent AML burden at this time point. By day 80 post-treatment, circulating AML cells reappeared in the reactivator pre-treated p53mt CAR-T cell group but remained significantly lower compared to untreated p53mt CAR-T cell-treated mice (Fig. 7C). These results indicate that while the p53 reactivator enhances the anti-AML function of p53mt CAR-T cells, their effects may wane over time. This decline could be attributed to progressive T-cell exhaustion, partial reversion of p53 activity, or downregulation of essential survival and metabolic pathways. Further studies are required to determine whether continuous or intermittent administration of the reactivator can sustain CAR-T cell function and improve long-term therapeutic outcomes.

Additionally, analysis of circulating CD3+ T cells revealed persistent T-cell circulation in both the reactivator pre-treated and untreated p53mt CAR-T cell groups. However, the reactivator pre-treated group exhibited a higher frequency of circulating T cells at both day 20 and day 80 (**Fig. 7D**), which correlated with improved mouse survival. This observation aligns with prior studies^21–23^ demonstrating that prolonged CAR-T cell persistence is associated with better therapeutic outcomes. Consistently, we observed that pre-treated p53mt CAR-T cells significantly extended mouse survival compared to untreated p53mt CAR-T cells, while no significant survival difference was observed between the reactivator pre-treated group and the control CAR-T cell-treated group (**Fig. 7E**). Collectively, these findings provide compelling evidence that the p53 reactivator restores the anti-tumor function of p53mt CAR-T cells, leading to survival benefits comparable to those of control CAR-T cells.

## Discussion

TP53 mutations, frequently observed in cancers, compromise the DNA binding capacity of p53 and diminish its tumor-suppressing abilities. In de novo acute myeloid leukemias (AML) and myelodysplastic syndromes (MDS), TP53 mutations are found in approximately 5-10% of cases ^24,25^, with increases to 30-40% in therapy-related or relapsed cases^26^. These mutations are significant prognostic factors in AML and MDS, invariably correlating with poor outcomes and resistance to standard chemotherapy ^27^. As a result, immunotherapeutic approaches are under investigation, including CD47-blocking therapies, immune checkpoint inhibitors, and T-cell and NK-cell-based cellular therapies. Despite variable success of these therapies in other malignancies, the clinical response of TP53-mutant AML, particularly cases with bi-allelic p53 or locus loss, has been dismal.

Currently, it is widely believed that the bone marrow microenvironment in leukemia is the primary mechanism behind the failure of immunotherapy. This belief has driven the development of numerous therapeutic strategies aimed at modifying the microenvironment, such as stromal-targeting agents^28^, hypoxia-modifying drugs^29^, and immune checkpoint inhibitors^30^. While these approaches have shown promise in preclinical models and select patient cohorts, they have largely failed to achieve durable responses in TP53-mutant AML. Our study suggests that an alternative mechanism, specifically the intrinsic dysfunction of TP53-mutant immune cells, may also contribute significantly to immune escape and therapy resistance. However, based on our observations of immune cell alterations in bone marrow samples from AML patients carrying TP53 mutations, our study proposes a new mechanism. Specifically, we suggest that TP53 mutations in immune cells induce their dysfunction, allowing AML cells to evade immune surveillance. To our knowledge, this is the first report linking TP53 mutations in T-cells to T-cell dysfunction.

Although some studies have detected TP53 mutations in T-cells from AML patients, they primarily identified these mutations as indicators of early pre-leukemic events. For instance, John Dick and his team reported^8^ that TP53 mutations in T-cells serve as markers for pre-leukemic mutations, without elucidating their functional impact. Similarly, Takahashi noted^9^ the presence of TP53 mutations in T-cells in the context of evolving pre-leukemic mutations and the development of AML. Another study ^31^published in Blood highlighted the persistence of TP53 mutations in T-cells but did not consider the potential impact on immune function. Collectively, these studies did not investigate whether TP53 mutations affect T-cell activity or contribute to therapeutic resistance. In contrast, our study addresses this critical gap by demonstrating that TP53-mutant T cells exhibit impaired cytotoxicity and increased exhaustion, thereby promoting immune evasion and AML progression. Notably, this contrasts with a recent report by Garcia et al.^32^, which showed that certain mutations derived from T cell malignancies—such as the CARD11–PIK3R3 fusion—can enhance CAR-T cell efficacy. Together, these findings underscore the context-dependent effects of somatic mutations in T cells, which may either hinder or augment anti-tumor immunity depending on the specific mutation and its downstream signaling pathways Utilizing advanced single-cell technologies, including scDNA sequencing integrated with surface antigen expression profiling, we conducted a comprehensive analysis of TP53 mutations across diverse immune cell populations at single-cell resolution. Subsequent validation via ddPCR confirmed the presence of TP53 mutations in multiple immune subsets, including T cells, NK cells, B cells, and monocytes. Since most immunotherapies focus on activating and restoring T-cell function, we concentrated our study on T-cells carrying TP53 mutations to understand their impact on immune cells. Our scDNA sequencing data revealed that TP53-mutant T-cells predominantly harbored monoallelic mutations and exhibited a distinct proteomic profile compared to their TP53 wild-type (WT) counterparts. Notably, these mutant T-cells demonstrated heightened proliferation, a characteristic frequently associated with p53 loss-of-function^33^. Given that the p53 protein functions as a tetramer, mutant p53 can bind to wild-type p53, potentially causing a dominant-negative effect^1,34,35^, which impairs the function of the remaining wild-type allele.

Due to technological constraints, directly assessing the function of p53 mutant T-cells derived from patients was not feasible. To address this limitation, we generated p53 mutant CAR-T cells from healthy donor T-cells, enabling a controlled investigation into the effects of mutant p53 expression in T-cells. This approach allowed us to evaluate the anti-tumor functionality of T-cells harboring mutant p53 against AML cell targets. Our experimental data demonstrated that p53 mutant CAR-T cells exhibited significantly enhanced proliferation, as evidenced by elevated expression of Ki67, CD69, and CD11b, compared to control CAR-T cells lacking mutant p53 expression. Furthermore, these p53 mutant CAR-T cells displayed an increased expression of exhaustion markers, including PD-1, TIM3, TIGIT, LAG3, and CD39, aligning with our scRNAseq analysis of T-cells from 24 AML patients. Specifically, T-cells from TP53-mutant patients consistently exhibited higher exhaustion marker expression. Cytotoxicity assays, conducted both in vitro and in vivo, further reinforced our hypothesis that mutant p53 compromises T-cell anti-tumor functionality.

To substantiate our hypothesis that mutant p53 drives T-cell dysfunction, we employed a targeted molecule, the p53-Y220C reactivator, which selectively restores the wild-type conformation of the p53-Y220C mutant. Although this compound remains under clinical investigation, we and others have demonstrated in preclinical studies its ability to reestablish wild-type p53 functionality^19,20^. Treatment of p53 mutant CAR-T cells with this reactivator resulted in a marked reduction in mutant p53 levels and a concomitant decrease in exhaustion marker expression in T-cells. Consequently, the anti-AML activity of p53 mutant CAR-T cells was significantly restored in both in vitro and in vivo models. These findings further support our hypothesis that mutant p53 actively contributes to T-cell dysfunction and that restoring its wild-type function can reverse this impairment.

Our study reveals a novel mechanism that may contribute to CAR-T therapy failure in AML. While previous research has focused on challenges such as the absence of ideal surface targets on AML cells and the suppressive tumor microenvironment, the role of mutations within the T-cells themselves has been largely overlooked. Our findings highlight TP53 mutations in T-cells from AML patients as a potential factor impairing T-cell anti-tumor function and leading to CAR-T cell therapy failure. Although TP53 mutations are present in only a subset of patient-derived T-cells, we observed that these mutant T-cells exhibited significantly enhanced proliferative capacity compared to their wild-type counterparts under in vitro culture conditions. This unchecked proliferation raises concerns that TP53-mutant T-cells could expand disproportionately within CAR-T products prior to infusion, potentially leading to an exhausted and dysfunctional phenotype. Given that T-cell exhaustion is a well-established contributor to reduced CAR-T efficacy, our findings suggest that the presence of mutant p53-expressing T-cells could directly compromise therapeutic outcomes. These results suggest that TP53 mutations not only serve as prognostic biomarkers but also actively drive immune dysfunction, a critical determinant of immunotherapy success. By demonstrating that mutant p53 can be selectively targeted to restore T-cell function, our research opens new avenues for therapeutic intervention. Collectively, our study provides compelling evidence that p53-mutant T-cells contribute to immune suppression in AML, and we establish proof-of-concept that restoring wild-type p53 activity in these cells can reverse their dysfunction and reinvigorate their anti-tumor capabilities. This concept is being under tested in a clinical trial of Rezatapopt (PC14586) in patients with MDS and AML carrying TP53-y220c mutations.

A key challenge moving forward is determining the precise origin of these mutations. One possibility is that TP53 mutations arise from clonal hematopoiesis of indeterminate potential (CHIP), a pre-malignant state in which hematopoietic stem cells acquire somatic mutations that expand over time. Alternatively, TP53 mutations may occur in hematopoietic progenitor cells prior to T-cell differentiation, leading to their persistence in T-cells and subsequent alterations in immune responses that contribute to AML progression. Clarifying the origin of these mutations is essential for understanding their role in AML pathogenesis and immune evasion and will aid in the development of targeted strategies to prevent the expansion of TP53-mutant immune cells and enhance immunotherapeutic efficacy.

## Data Availability

scRNA-seq data are publicly available in Gene Expression Omnibus (GEO) repository. The CyTOF data are publicly available in the Flow Repository.

## Acknowledgement

Supported by the NIH/NCI under award number P30 CA016672 and used MDACC Cancer Center Support Grant (CCSG) shared resources.

