## Supplementary material for "Somatic TP53 Mutations Drive T and NK Cell Dysfunction in AML and Can be Rescued by Reactivating Wild Type p53": methods with supplementary.pdf

#### Single Cell DNA+Protein Sample Staining and Preparation

Samples with at least 70% viability were stained with the Total-Seq™-D Human Heme Oncology Cocktail. This cocktail contains 45 oligo-conjugated antibodies targeting various surface markers relevant to heme oncology. Following staining, the cells were washed to remove unbound antibodies and resuspended in a suitable buffer. Single-cell suspension, encapsulation, and barcoding were performed according to the manufacturer's instruction. The encapsulation process involved using the Chromium system (10x Genomics), which enables the generation of gel beads in emulsion (GEMs) containing barcoded oligonucleotides that uniquely label each cell.

#### scDNA Library Preparation and Sequencing

For scDNA library preparation, we utilized a validated custom panel (Morita et al. 2020) consisting of 279 amplicons covering recurrent mutations in 37 genes in AML. The panel was designed to target regions frequently mutated in AML, including TP53, FLT3, NPM1, and CEBPA, among others. The library was prepared using the Mission Bio Tapestri platform, which enables simultaneous genotyping and phenotyping of single cells. The prepared libraries were then sequenced using an Illumina sequencer, following which the raw sequencing data were demultiplexed, aligned, and processed for downstream analysis.

#### scRNA-seq

Single-cell RNA sequencing (scRNA-seq) was performed on a cohort of 29 samples, including 3 normal bone marrow samples and 26 AML samples collected at the time of diagnosis (19 *TP53*-mut, 7 *TP53*-wt). Cells were dissociated and prepared for scRNA-seq using a standard protocol, ensuring high viability and minimal cell loss. The single-cell libraries were generated using the Chromium Single Cell 3' Reagent Kits v3 (10x Genomics) according to the manufacturer’s protocol, followed by sequencing on an Illumina platform. Data processing and quality control were carried out using the Cell Ranger software (10x Genomics). High-quality cells with a minimum of 500 genes and less than 10% mitochondrial RNA content were retained for downstream analysis. The

Scanpy package (version 1.7.2) was used for data normalization, scaling, and identification of highly variable genes. Cells from all samples were clustered using the Leiden algorithm, and the cell identities were annotated with the CellTypeST tool. This annotation leveraged known marker genes to assign cell types to the clusters, ensuring accurate classification of hematopoietic and leukemic subpopulations. The abundance of different cell populations, including hematopoietic stem and progenitor cells (HSPCs), erythroid cells, and various myeloid lineages, was quantified and compared across samples.

**Mass cytometry data analysis**

**Sample Barcoding, CyTOF Staining, and Data Acquisition**

Samples were barcoded using the Cell-ID 20-Plex Pd barcoding kit (Standard BioTools, San Francisco, CA), which employs triple combinations of six distinct palladium (Pd) isotopes. Cells were fixed, washed once in 1X barcode perm buffer, and resuspended in 800 µL barcode perm buffer. Barcodes were thawed at room temperature, quick-spun, and resuspended in 100 µL barcode perm buffer, then mixed thoroughly with the corresponding samples and incubated at room temperature for 30 minutes. Following incubation, samples were washed three times with cell staining buffer (0.5% bovine serum albumin [BSA] in PBS), resuspended in 100 µL PBS, and pooled into a single tube. Prior to staining, cells were counted, centrifuged, and resuspended in cell staining buffer.

For CyTOF staining and data acquisition, cells were labeled with 5-Iodo-2'-deoxyuridine (IdU) (Acros Organics) to identify cells in the S-phase of the cell cycle. Cells were incubated with 10 µM IdU for 30 minutes at 37°C and 5% $CO_2$, followed by two washes with cell staining buffer (CSB). Pooled samples were incubated with Human TruStain FcX™ solution (BioLegend) for 10 minutes to block Fc receptors, then stained for 30 minutes at room temperature with a freshly prepared antibody mixture targeting cell surface markers. After incubation, cells were washed twice with CSB, fixed in 1.6% paraformaldehyde (PFA) for 10 minutes at room temperature, and permeabilized with 90% methanol at -20°C for 1 hour. Cells were subsequently washed twice in CSB, stained with intracellular antibodies for 30 minutes at 4°C, and resuspended in intercalator solution for overnight incubation at 4°C. Before acquisition, cells were washed twice in CSB, once with double-distilled water, and analyzed using a Helios mass cytometer.

**Data Analysis**

Deconvolution of pooled sample sets was performed using the Premessa R package or Debarcoder software (Standard BioTools, San Francisco, CA), followed by data clean-up in FlowJo (version 10.8.1). Single, live cells were selected by excluding calibration beads, gating on singlets based on DNA content and event length, and removing dead cells by selecting populations with low cisplatin uptake. High-dimensional data analysis was conducted using the Omiq.ai platform, with Uniform Manifold Approximation and Projection (UMAP) applied for dimensionality reduction. Fish plots and violin plots were generated using R. All datasets were tested for normal distribution prior to statistical analysis, with parametric variables analyzed using the t-test, and nonparametric datasets assessed using the Mann-Whitney U test.

**CAR-T cells re-challenge experiment**

Control-CAR-T cells and p53mut-CAR-T cells were co-cultured with Molm13 cells at 1:1 ratio(1X10^6 cells) in 24 well plate without any cytokine in the RPMI-1640 medium (Gibco, CAT,11875093) with 10% fetal bovine serum(Gibco, CAT, A5670801) in 37C, 5% CO2 incubator, and re-challenged with 1X10^6 cells Molm13 cells/well every other day until day14. Analysis of T cells phenotype was analyzed by cyTOF at different timepoints.

**Incucyte Live-Cell Cytotoxicity assay**

CAR-T cells were co-cultured with tumor cells at an effector-to-target (E:T) ratio of 1:5. Target cells were labeled with Vybrant DyeCycle Ruby Stain (ThermoFisher) to facilitate identification. To detect apoptosis, the Incucyte Caspase-3/7 Green Apoptosis Assay Reagent (Sartorius) was added to each well.

Real-time imaging was conducted over a 72-hour period using the Incucyte Live-Cell Analysis System, which continuously captured images and quantified apoptosis. The system tracked apoptotic cells (green fluorescence) and target cells (red fluorescence). The percentage of caspase-3/7 activation was calculated as the proportion of dual-positive (green and red) target cells relative to the total red-labeled tumor cells, providing a dynamic measure of tumor cell apoptosis.

#### Single-Cell Cytotoxicity Assay

Time-lapse imaging microscopy in Nanowell grids (TIMING)[1] was utilized to assess CAR-T cell cytotoxicity at a single-cell resolution, as previously described. Briefly, CAR-T cells and Molm13 target cells were differentially labeled with lipophilic PKH dyes and loaded onto Nanowell arrays. The arrays were incubated in culture medium pre-mixed with Annexin V (BD Biosciences) to detect apoptotic events in real time.

Live-cell imaging was performed for 5–6 hours using a Carl Zeiss Axio Observer microscope, equipped with a Hamamatsu Orca-Flash sCMOS camera and a ×20, 0.8 numerical aperture objective. Approximately 5,000 individual wells were imaged and analyzed using a custom in-house algorithm for cell tracking and segmentation.

#### Chimeric antigen receptor lentiviral vector production

Lentiviral vectors encoding the anti-CD123 single-chain variable fragment (scFv), with or without p53-Y220C, were synthesized and subcloned into the pCCL backbone. The construct was designed in-frame with sequences encoding the IgG1 hinge region, the CD28 transmembrane domain, and CD3ζ intracellular signaling domains, under the control of the EF1α promoter, as illustrated in Supplementary Fig. 1. Lentiviral supernatant was generated, collected, and used for T-cell transduction, following previously described protocols [2].

#### Western Blot Analysis

Western blotting was performed as previously described[3] using the Odyssey Infrared Imaging System (LI-COR Biosciences, Lincoln, NE) for signal detection and Odyssey software version 3.0 for quantification. Protein lysates were resolved on SDS-PAGE gels and transferred to PVDF membranes. Membranes were blocked and incubated with primary antibodies, including anti-p53 (DO-1; Santa Cruz Biotechnology, #sc-126), anti-MDM2 (D-12; Santa Cruz Biotechnology, #sc-5304), and anti-p21 (12D1; Cell Signaling Technology, #2947). GAPDH (14C10; Cell Signaling Technology, #2118) served as a loading control. After incubation with appropriate IRDye-labeled secondary antibodies, signals were visualized using the Odyssey system, and band intensities were quantified using Odyssey software.

**In vivo experiment**

PDX AML cells were obtained from an AML patient resistant to Ven and DAC therapy and were established in our laboratory[3] . Post-treatment, the AML cells harbored FLT3-ITD (two ITDs with allele ratios of 0.235 and 0.119), GATA2 (p.R362*), and NRAS (p.G12D, <2% variant allele frequency) mutations. A total of 1–2 million PDX AML cells were injected intravenously into 6-week-old male and female NSG mice. Engraftment was confirmed via flow cytometry when human CD45 (Pacific Blue, BioLegend, #304029) and CD33 (PE, BioLegend, #366608) double-positive cells exceeded 1% of the combined mouse CD45-negative and human CD45-positive populations in peripheral blood (PB) samples. Mice were randomized based on circulating leukemia burden and body weight.

One day after confirming engraftment, $3 \times 10^6$ T cells/mouse were administered intravenously. Treatment continued until all mice in each group became moribund. Leukemia burden was monitored weekly by flow cytometry, measuring human CD45 and CD33 double-positive cells in PB.


1 Liadi, I., Roszik, J., Romain, G., Cooper, L. J. & Varadarajan, N. Quantitative high-throughput single-cell cytotoxicity assay for T cells. *J Vis Exp*, e50058 (2013). https://doi.org/10.3791/50058

2 Mu-Mosley, H. *et al.* Transgenic Expression of IL15 Retains CD123-Redirected T Cells in a Less Differentiated State Resulting in Improved Anti-AML Activity in Autologous AML PDX Models. *Front Immunol* **13**, 880108 (2022). https://doi.org/10.3389/fimmu.2022.880108

3 Carter, B. Z. *et al.* Targeting MCL-1 dysregulates cell metabolism and leukemia-stroma interactions and resensitizes acute myeloid leukemia to BCL-2 inhibition. *Haematologica* **107**, 58-76 (2022). https://doi.org/10.3324/haematol.2020.260331

### Supplementary Table 1. exhaustion-associated markers

| |
|---|
| PDCD1 |
| CTLA4 |
| HAVCR2 |
| LAG3 |
| TIGIT |
| CD244 |
| EOMES |
| TOX |
| ENTPD1 |
| NT5E |
| BATF |
| CD160 |
| KLRG1 |
| VSIR |
| BTLA |
| NFATC1 |
| IL10 |
| TGFB1 |
| CD200 |
| CD96 |
| LGALS9 |

Supplementary Table 2. T cell panel for CyTOF

| T cell panel | | | |
| --- | --- | --- | --- |
| 89 | Barcode | 154 | pMCL1 |
| 102 | Barcode | 155 | TP53 WT |
| 103 | Barcode | 156 | TIM3 |
| 104 | Barcode | 157 | pMCL1 |
| 105 | Barcode | 158 | 2B4 |
| 106 | Barcode | 159 | ICOS |
| 108 | Barcode | 160 | PD1 |
| 110 | Barcode | 161 | CXCR3 |
| 111 | Barcode | 162 | total P53 |
| 112 | Barcode | 163 | CTLA4 |
| 113 | CD8 | 164 | CD96 |
| 114 | | 165 | CD25 |
| 115 | CD3 | 166 | LAG3 |
| 116 | H2AX | 167 | Perforin |
| 127 | IDU | 168 | CD160 |
| 139 | CD36 | 169 | **p53** |
| 140 | | 170 | CD39 |
| 141 | CD38 | 171 | CD161 |
| 142 | CD27 | 172 | KLRG2 |
| 143 | CD56 | 173 | Bax |
| 144 | BCL2 | 174 | GITR |
| 145 | NKG2A | 175 | SATB1 |
| 146 | CD49F | 176 | TOX |
| 147 | CD127 | 194 | LD |
| 148 | CCR2 | 195 | CD57 |
| 149 | CCR4 | 196 | CD4 |
| 150 | LDHB | 198 | HLA DR |
| 151 | P53 | 208 | CD97 |
| 152 | TIGIT | 209 | CD11b |
| 153 | CD45RA | | |

Supplementary Figure 1.

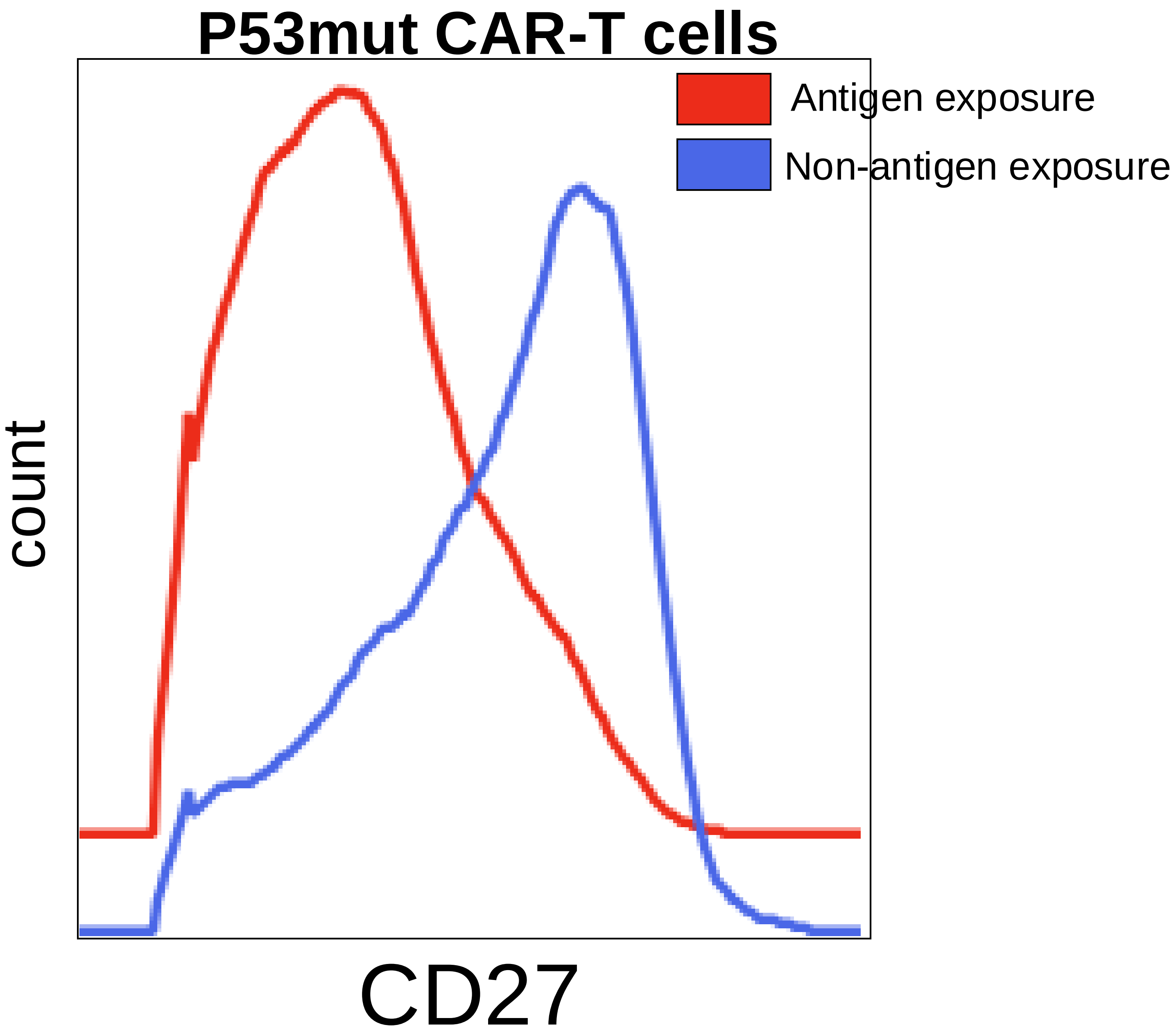

Supplementary Figure 2.

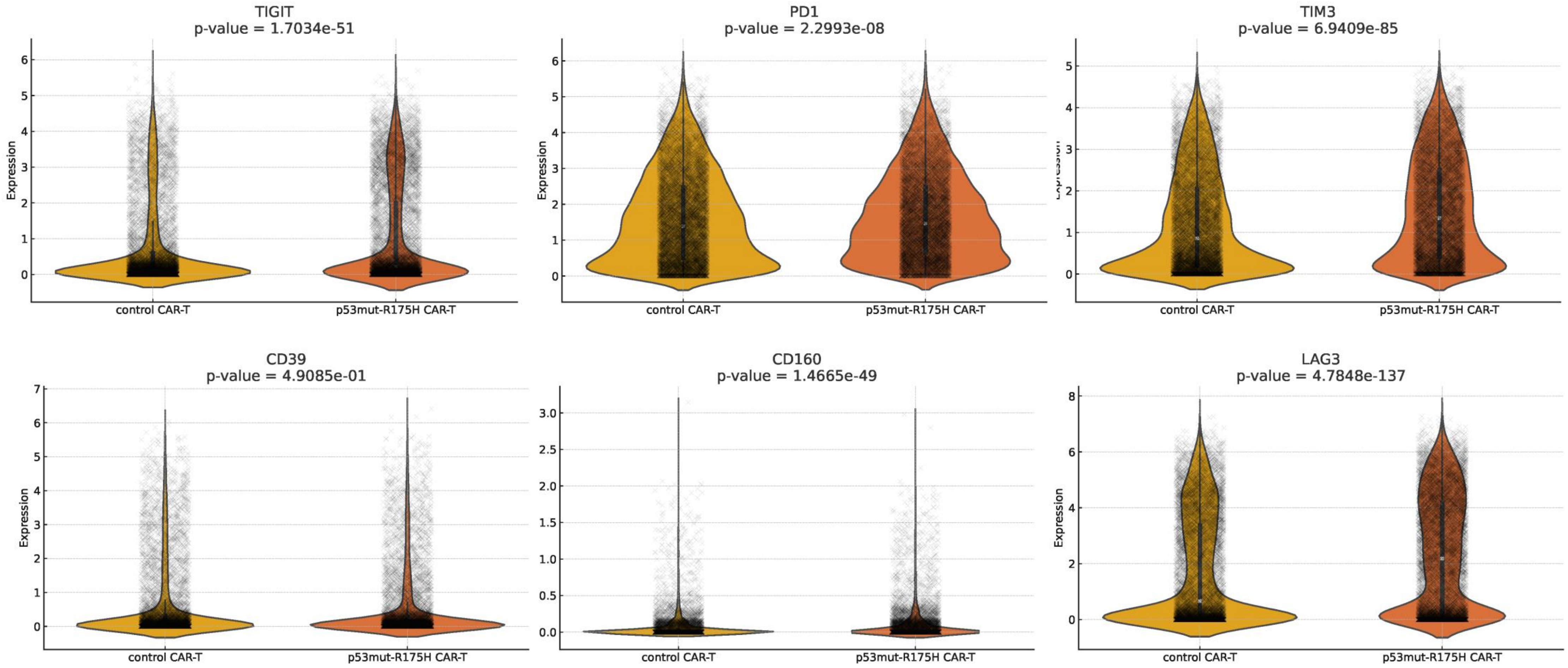

mouse T cells phenotype

Supplementary Figure 3.

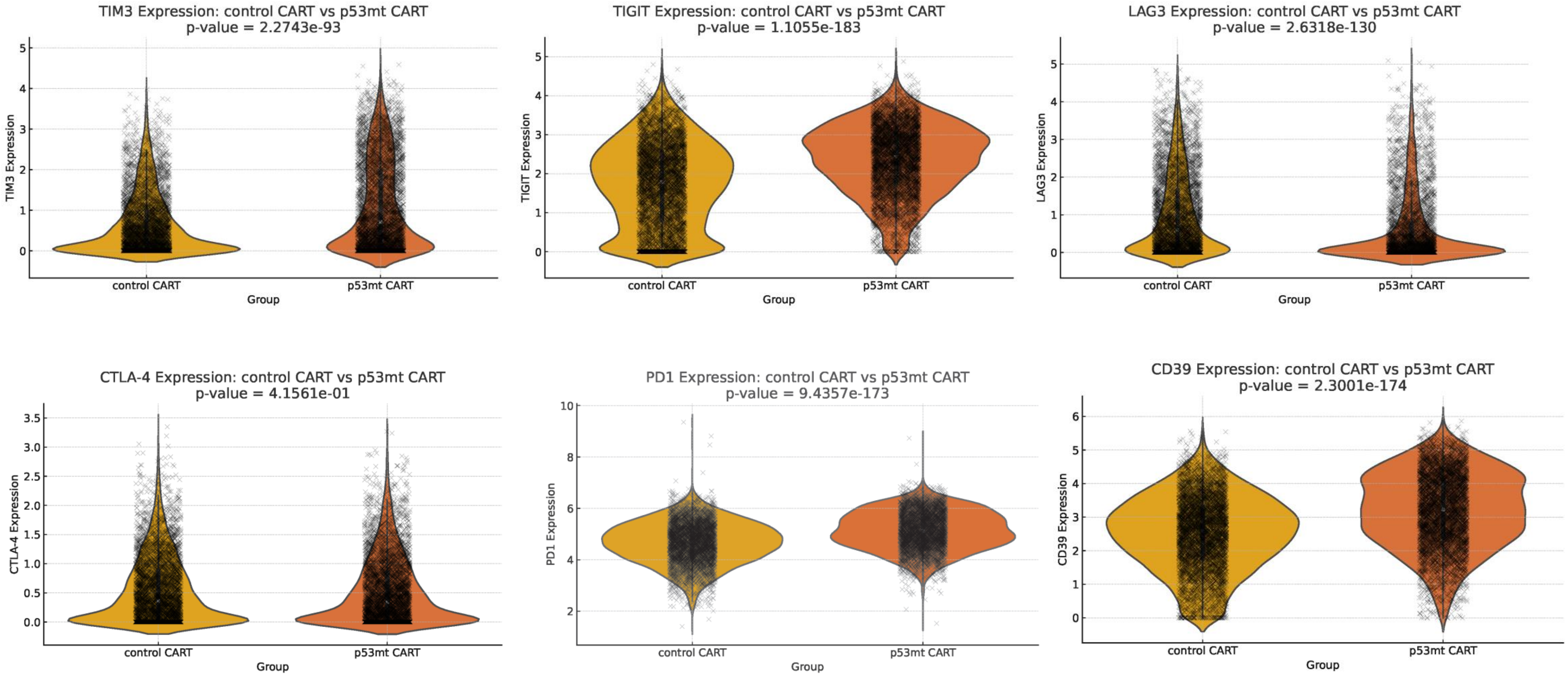

**Supplementary figure 4**

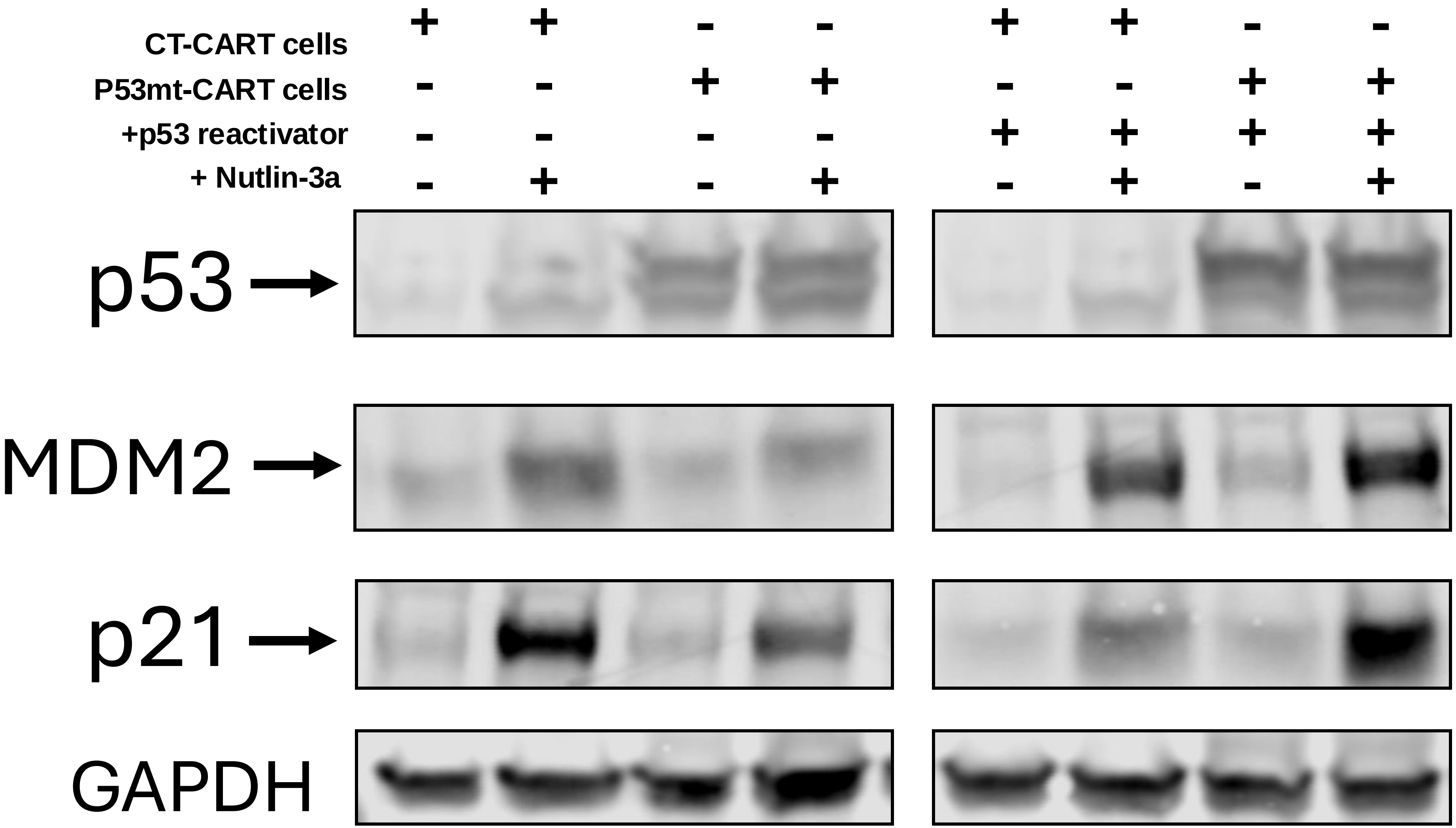
